# Characterization of coordinated growth in macrodactyly caused by somatic mosaic activating mutations in *PIK3CA*

**DOI:** 10.1101/2022.06.07.22275709

**Authors:** Catherine McNamara, Jennifer Lanni, Jake Daane, Laura Nuzzi, David Peal, Matthew P. Harris, Brian Labow

## Abstract

Localized somatic overgrowth disorders that occur during development can be debilitating, and most often require surgical intervention. Although underlying genetic changes associated with overgrowth have been identified in the majority of cases, the cause of the dysregulated growth and its presentation is unknown. Here we detail current work on a specific overgrowth disorder, macrodactyly, in which overgrowth is localized and shows integration with developmental patterning of the limb, providing coordination of the resulting overgrowth structure. We provide clinical analysis of presentation of macrodactyly in a cohort of patients and provide experimental evidence for nerve and vascular-biased regulation of growth. We provide the first animal model that recapitulates macrodactyly and provide evidence that genetic modifiers may underlie the development of this disorder. The unique presentation of macrodactyly provides a framework to identify the causes and regulatory activities that shape hyperplastic signals that lead to integrated patterning in overgrowth. Use of our experimental model suggests potential for genetic modifiers as important for the particular presentation of this disorder over other PIK3CA-related growth disorders.

## INTRODUCTION

Dysregulated growth of somatic tissue during development characterizes an enigmatic class of disorders with varied phenotypic presentations that often require surgical intervention for maintenance of care and quality of life. A subset of these conditions, known as *PIK3CA*-Related Overgrowth Spectrum (PROS) disorders, harbor somatic, activating *PIK3CA* mutations and encompass anomalies such as macrodactyly, congenital lipomatous overgrowth, vascular malformations, epidermal naevi, scoliosis/skeletal and spinal syndrome (CLOVES), and Klippel-Trenaunay (KT) syndrome (Keppler-Noreuil et al., 2015; Martinez-Lopez et al., 2019). Under normal growth conditions, *PIK3CA* is a critical member of the phosphoinositide 3-kinase (PI3K) pathway, which is an essential regulator of cell proliferation, metabolism, and growth through the activity of downstream effectors such as AKT and mTOR. Although specifically linked to these somatic overgrowth disorders in development, *PIK3CA* is frequently altered in cancers as well and often contributes to uncontrolled, hyperplastic or neoplastic overgrowth (Samuels et al., 2004). Recent advances in detecting rare somatic mutations in clinical samples have enabled identification of common *PIK3CA* driver mutation hotspots (such as H1047R, E545K, and E542K) that are shared among overgrowth syndromes with different regional growth characteristics (Fruman et al., 2017; Ren et al., 2021; Yeung et al., 2017).

Despite this common genotype, phenotypic presentations among the PROS disorders can be starkly unique(Keppler-Noreuil et al., 2015). Of interest in our study is macrodactyly, which presents as coordinated, patterned overgrowth that unilaterally affects multiple digits of the hand or foot (**Figure 1**) (Cerrato et al., 2013; Flatt, 1994). The presentation of macrodactyly often is biased to nerve territories, associating with radial territories. This bias has led to hypotheses concerning nervous tissue regulated control of the overgrowth. In contrast to the presentation of macrodactyly, patients with CLOVES or KT more often present with uncoordinated, non-patterned overgrowth. A limited number of studies have reported *PIK3CA* mutations in macrodactyly; however, the rarity of this condition has resulted in little information regarding the *PIK3CA*-specific role in disease progression or in macrodactyly’s unique phenotype and disease progression (Bornstein et al., 2014; Cappuccio et al., 2017; Hucthagowder et al., 2017; Keppler-Noreuil et al., 2014; Rios et al., 2013; Tripolszki et al., 2016; Wu et al., 2020b; Wu et al., 2018; Yeung et al., 2017).

**Figure 1.**
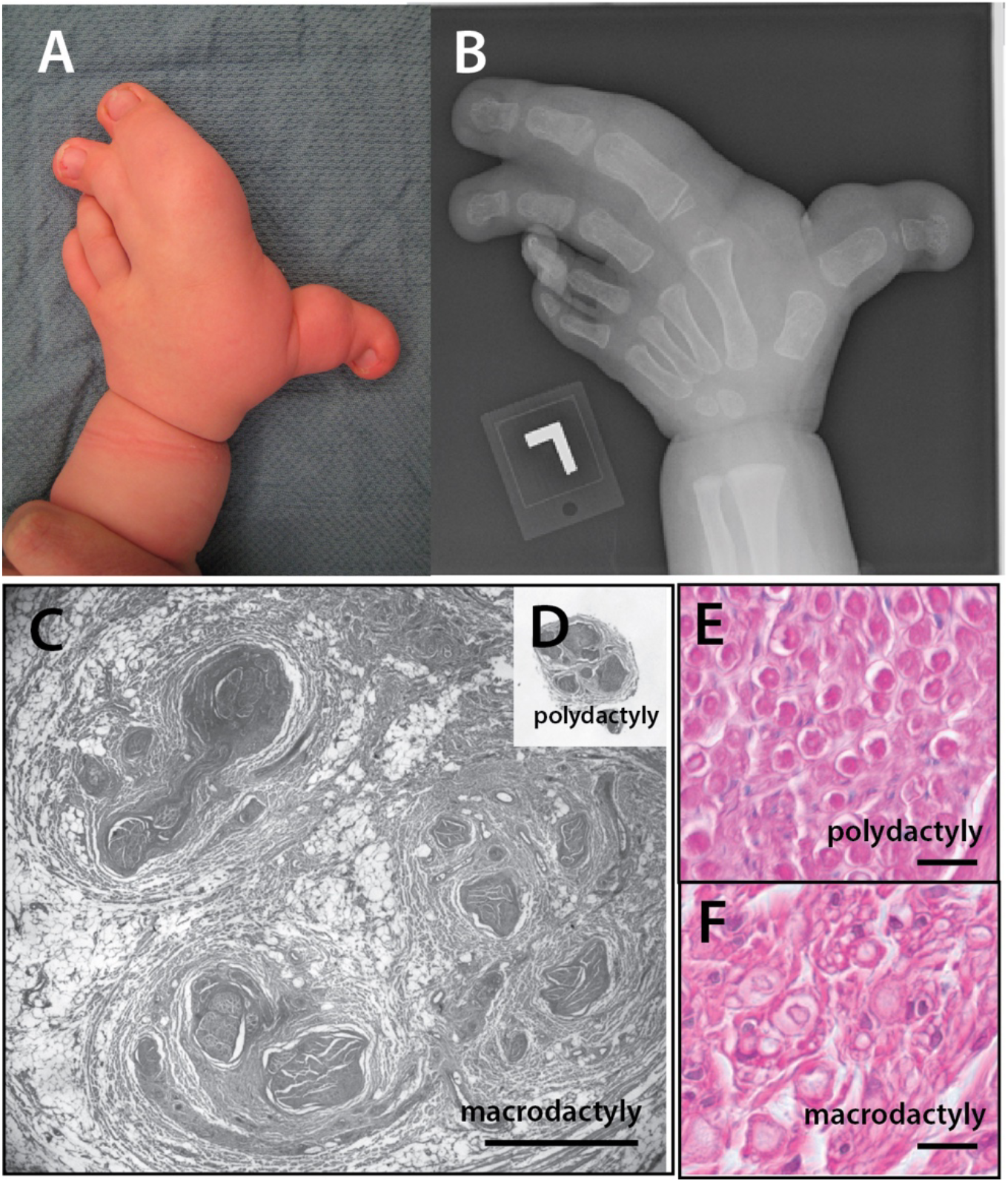
Patterned overgrowth in digital elements in macrodactyly. **A, B** Overgrowth in macrodactyly is characterized by coordinated, patterned overgrowth of the skeleton and associated tissues of the limbs. Such coordinated overgrowth predominantly occurs in the hand/foot with rare proximal effects extending into the forearm. **C-F** Macrodactyly is often associated with hypertrophy of nerves and fibroadipose tissue (C, cross section of digital nerve from macrodactyly patient compared to D, polydactyly digital nerve at same magnification; bar, 1mm). The hypertrophy of the nerve in macrodactyly (F) can be associated with abnormal growth of the glia and fibroadipose tissue when compared to polydactyly (E; bar, 20um).

Given the commonality of PIK3CA gain-of-function mutations within growth disorders of PROS, it is generally assumed PIK3CA activation in somatic clones is sufficient to drive the segmental overgrowth and vascular phenotypes. Prior models of PIK3CA somatic activating clones in mice have shown that this mutation is indeed sufficient to elicit many of the vascular pathologies (Hare et al., 2015) as well as overgrowth of skeletal muscle, megalocephaly and organomegaly (Kinross et al., 2015; Mitchell and Phillips, 2019) associated with these disorders. However, experimental studies to date have not been able to mirror the coordinated overgrowth phenotype of the distal limb observed in macrodactyly. The inability of prior models of activated *PIK3CA* to mirror growth observed in macrodactyly suggests added, complex regulation of PIK3CA signaling within macrodactyly may provide critical insight into control of these disorders.

In our study, we aim to understand the contribution that somatic mutations make to the developmental and phenotypic presentation of macrodactyly. To do so, we closely analyze mutational status from affected tissue from a robust cohort of patients diagnosed with macrodactyly as well as their demographic and clinical data. We assess the genotype of a broad set of growth related genes within these tissues and highlight potential genetic modifiers of PIK3CA altered function. Using the diversity of patient material, we address the expression of growth related genes within overgrown tissue and characterize trends in gross anatomy. We further extend these analyses and define a distinct genetic model to test *PI3KCA*-mediated overgrowth during development. Our data highlight an actionable experimental model in the zebrafish to understand genetic and anatomical regulation of coordinated growth as seen in macrodactyly, and how it might be leveraged to rein in progressive or neoplastic growth of other PROS conditions.

## RESULTS

Macrodactyly is a unique growth disorder caused by activation of PIK3 signaling that unlike other PROS disorders such as CLOVES and/or Klippel-Trenaunay syndromes, leads to localized, patterned overgrowth of the distal limbs. The coordination of growth makes macrodactyly a potentially unique overgrowth disorder that may reveal genetic or anatomical means of shaping or controlling otherwise disorganized overgrowth. Macrodactyly shows prominent bias to pre-axial or postaxial ‘territories’, as well as hypertrophied palmar/plantar surfaces suggesting involvement of sensory nerves in its etiology. Although early work has tied macrodactyly to known gain-of-function somatic mutations in PIK3CA, modeling of this disorder has not shown that this driver mutation is sufficient to cause the coordinated limb overgrowth observed in patients.

We used a robust cohort of patient samples to enable an assessment of mutation within other growth regulators within macrodactyly. Through a systematic analysis of macrodactyly patients seen at Boston Children’s Hospital we were able to analyze tissue specimens from thirty-six patients. The cohort had a median age of 1.5 years at initial surgery (minimum: 0.5, maximum: 13.8, interquartile range (IQR): 1.2)(**Supplementary Table 1, Table 1**). Half of patients were male, and the majority of patients were self-identified as white, non-Hispanic (58.3%; 21/36) with scattered representation of other racial groups. Nearly 92% of patients experienced overgrowth of multiple digits within the affected extremity, with a median of 2.0 digits affected (minimum: 1.0, maximum: 3.0, IQR: 1.0). Approximately 89% of patients presented with isolated macrodactyly, and greater than half of all patients (52.8%) were classified as having lipomatous macrodactyly. The left extremity was affected in 69.4% of patients, and half of affected patient samples (50.0%) were from the hand. The most commonly affected nerve territory was preaxial (47.2%), followed closely by central (41.7%), and then postaxial (11.1%) territories (**Table 1**).

**Table 1.** Summary of macrodactyly cohort and presentation.

| **Characteristics** | **Macrodactyly cohort** |
| --- | --- |
| **Sex** |  |
| Male | 18 (50%) |
| Female | 18 (50%) |
| **Race** |  |
| White | 21 (58.3%) |
| Other | 5 (13.9%) |
| Unknown | 4 (11.1%) |
| Hispanic | 3 (8.3%) |
| Black or African-American | 2 (5.6%) |
| Asian | 1 (2.8%) |
| **Median (IQR) age at initial surgery, years** | 1.5 (1.2) |
| **Type of macrodactyly** |  |
| Nerve-territory oriented | 17 (47.2) |
| Lipomatous | 19 (52.8) |
| **Macrodactyly presentation** |  |
| Isolated | 32 (88.9) |
| Secondary | 4 (11.1) |
| **Affected extremity** |  |
| Hand | 18 (50.0%) |
| Foot | 18 (50.0%) |
| **Affected side** |  |
| Left | 25 (69.4%) |
| Right | 11 (30.6%) |
| **Multiple digits affected** | 33 (91.7%) |
| Median (IQR) number of digits affected | 2.0 (1.0) |
| **Nerve Territory** |  |
| Preaxial | 17 (47.2%) |
| Central | 15 (41.7%) |
| Postaxial | 4 (11.1%) |
| **TOTAL** | **36** |

### Targeted capture approaches identify rare, low penetrant somatic clones

To provide increased coverage needed to detect low frequency somatic mutations within tissues, we used a targeted capture approach to mine the genetic variability of overgrown tissue. Given prior work linking overgrowth disorders such as Proteus, CLOVES and Macrodactyly to PIK3 and AKT signaling, we included oligo baits targeted to these genes. In addition, we included several genes that were identified as causing coordinated overgrowth of zebrafish fins when their function is altered (Daane et al., 2021; Lanni et al., 2019; Perathoner et al., 2014) (**Supplementary Table 1**). We sequenced resected tissue from localized digital outgrowths and directly compared unique, non-reference variants observed when compared with similar analysis from blood samples from the same patients. In this manner, we could detect somatic variants present at low frequency unique to the affected tissue as putative causative somatic changes associating with macrodactyly presentation.

Targeted sequencing showed efficient capture and provided an average of 84-95% fold coverage of loci included on the array, from >2 and >10 reads per locus, respectively per patient (**Supplementary Table 2**). Similar to analysis from previous studies of macrodactyly, as well as CLOVES and Klippel-Trenaunay syndromes, one of the frequent somatic mutations identified across individuals unique to affected tissue were nonsynonymous mutations in *PIK3CA* (**Table 2;** (Castel et al., 2016; Castillo et al., 2016; Kuentz et al., 2017; Kurek et al., 2012; Peyre et al., 2021; Rios et al., 2013; Rodriguez-Laguna et al., 2019; Tian et al., 2020; Wu et al., 2018; Yeung et al., 2017). The identified variants within *PIK3CA* are common in overgrowth disorders and define PROS. Additionally, these mutations are often found in human cancers suggesting they underlie growth potential in different contexts. Overall, known mutations in *PIK3CA* were found in approximately 89% of patients (32/36) (**Table 1**). Among these patients, mutations at H1047R were the most common, occurring in 66.7% (24/36) of patients. Much less common were mutations at E545K (*N*= 3, 8.3%), and E542K and H1046L (*N*=2, 5.6%, both). Mutations in C420R were rare, occurring in only one patient (2.8%). Overall *PIK3CA* mutation status did not track with anatomical location, age, sex, or race (*p* > 0.05, all).

**Table 2.**
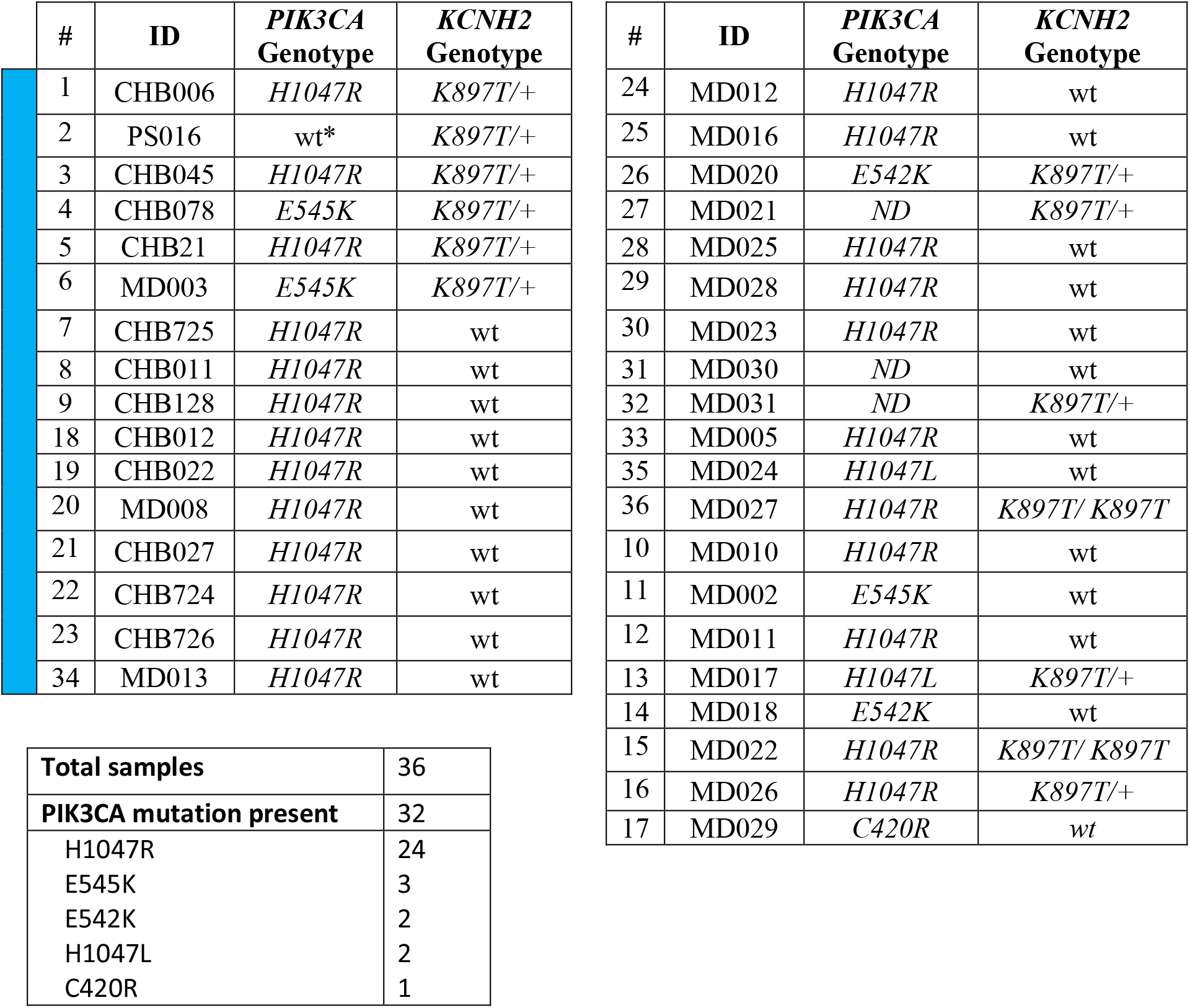
Genotype of *PIK3CA* and *KCNH2* in patient cohort. Blue bar – samples in which targeted sequence carried out. Right table genotyped for PIK3CA and KCNH2 only. *PIK3CG S442Y detected;

| # | ID | *PIK3CA*<br>Genotype | *KCNH2*<br>Genotype |
| --- | --- | --- | --- |
| 1 | CHB006 | *H1047R* | *K897T/+* |
| 2 | PS016 | wt* | *K897T/+* |
| 3 | CHB045 | *H1047R* | *K897T/+* |
| 4 | CHB078 | *E545K* | *K897T/+* |
| 5 | CHB21 | *H1047R* | *K897T/+* |
| 6 | MD003 | *E545K* | *K897T/+* |
| 7 | CHB725 | *H1047R* | wt |
| 8 | CHB011 | *H1047R* | wt |
| 9 | CHB128 | *H1047R* | wt |
| 18 | CHB012 | *H1047R* | wt |
| 19 | CHB022 | *H1047R* | wt |
| 20 | MD008 | *H1047R* | wt |
| 21 | CHB027 | *H1047R* | wt |
| 22 | CHB724 | *H1047R* | wt |
| 23 | CHB726 | *H1047R* | wt |
| 34 | MD013 | *H1047R* | wt |

|  |  |
| --- | --- |
| **Total samples** | 36 |
| ***PIK3CA* mutation present** | 32 |
| H1047R | 24 |
| E545K | 3 |
| E542K | 2 |
| H1047L | 2 |
| C420R | 1 |

| # | ID | *PIK3CA*<br>Genotype | *KCNH2*<br>Genotype |
| --- | --- | --- | --- |
| 24 | MD012 | *H1047R* | wt |
| 25 | MD016 | *H1047R* | wt |
| 26 | MD020 | *E542K* | *K897T/+* |
| 27 | MD021 | *ND* | *K897T/+* |
| 28 | MD025 | *H1047R* | wt |
| 29 | MD028 | *H1047R* | wt |
| 30 | MD023 | *H1047R* | wt |
| 31 | MD030 | *ND* | wt |
| 32 | MD031 | *ND* | *K897T/+* |
| 33 | MD005 | *H1047R* | wt |
| 35 | MD024 | *H1047L* | wt |
| 36 | MD027 | *H1047R* | *K897T/ K897T* |
| 10 | MD010 | *H1047R* | wt |
| 11 | MD002 | *E545K* | wt |
| 12 | MD011 | *H1047R* | wt |
| 13 | MD017 | *H1047L* | *K897T/+* |
| 14 | MD018 | *E542K* | wt |
| 15 | MD022 | *H1047R* | *K897T/ K897T* |
| 16 | MD026 | *H1047R* | *K897T/+* |
| 17 | MD029 | *C420R* | wt |

Interesting, wildtype *PIK3CA* was also found in one patient suggesting another etiology. This one patient was also found with somatic mutation in *PIK3CG*, encoding P100-KD catalytic subunit of PI3Kγ. This mutation is also common among our screened macrodactyly cohort (5 out of 17 patients), and was present in individuals also having *PIK3CA* H1047R mutations (3/17). The S442 site is predicted to act as a phosphorylation site (www.phosphonet.ca) and the S442Y mutation is predicted to be potentially deleterious (PROVEAN, -0.931 (neutral); Polyphen, 0.984 (damaging)) suggesting potential function of this mutation. Somatic mutations in *PIK3CG* have not been previously identified in PROS patients, however this variant can be found in human population at low frequency (11%, gnomad). Other nonsynonymous changes unique to affected tissue were found in several genes known to regulate growth (**Supplementary Table 3**). A missense mutation S299L was found in *NFATC1* in one patient and was further confirmed by Sanger sequencing suggesting this mutation was evident in the overgrown tissue. NFAT is a key regulator of transcription in response to calcium signaling in cells and is essential for normal development (Horsley and Pavlath, 2002). The S299L mutation is not predicted to be particularly deleterious (PROVEAN, -2.58; Polyphen, 0.31). No mTOR or Akt mutations were identified in this cohort of patients.

As the mutations in macrodactyly are mosaic, different tissues can have relative cellular contributions with altered gene function. Given the aberrant size of the nerve in many cases of macrodactyly (**Figure 1**), we were in some instances able to dissociate nerve from associated adipose tissue. From these samples, we used bulk sanger sequencing to approximate the level of variant mutation alleles within the sample population. Results show that compared to general adipose tissue, nerve contains a high proportion of altered *PIK3CA* transcripts (**Figure 2B**). This supports the potential etiology of nerve-directed function underlying this disorder (e.g.(Moore, 1942)).

**Figure 2.**
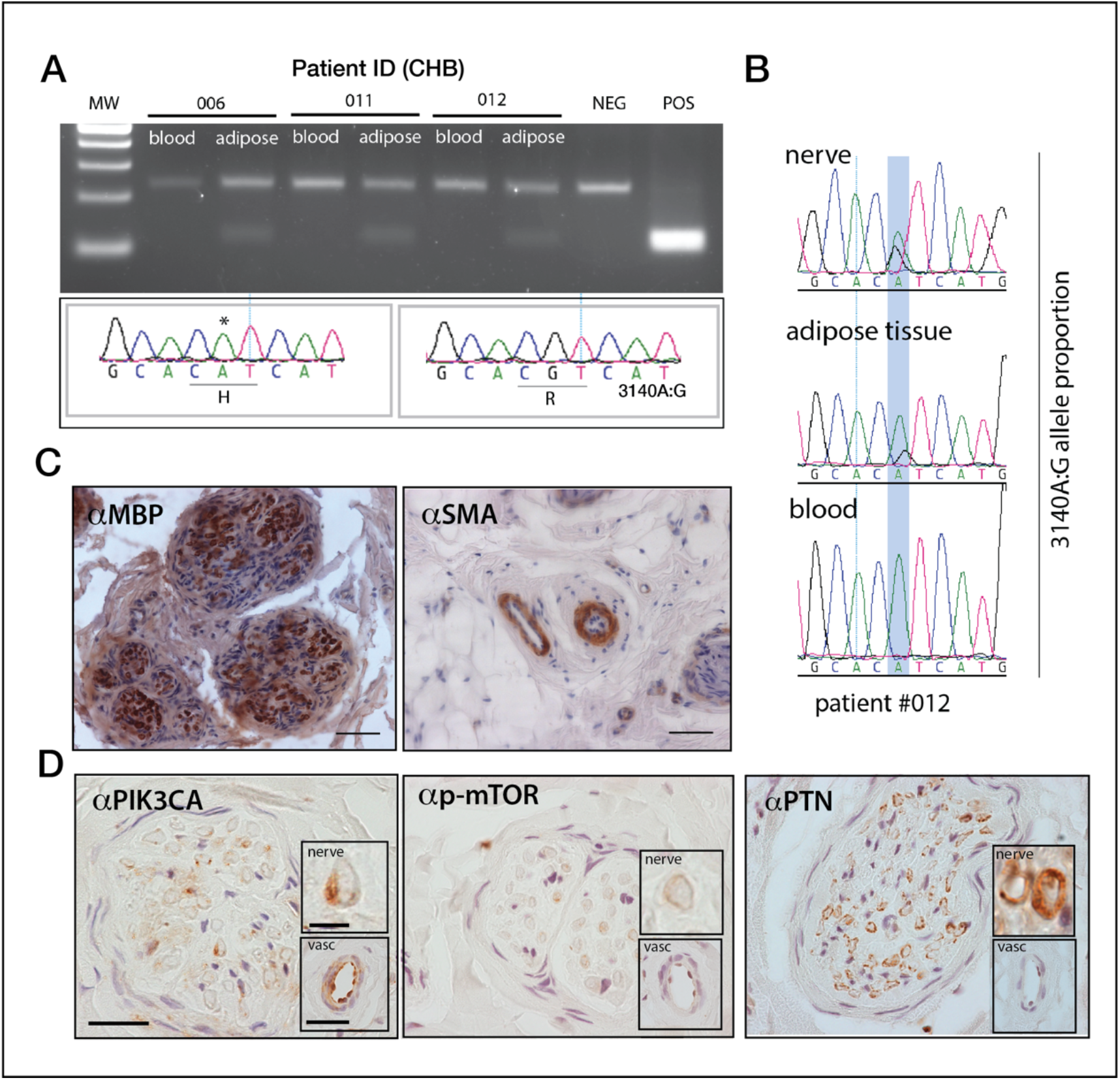
A common residue in PIK3CA is frequently mutated in macrodactyly as well as other growth disorders. **A)**. Detection of c3140A->G mutation in *PIK3CA* by HpyCH4IV enzymatic digest of PCR products of *PIK3CA* Exon 21 from affected tissue of macrodactyly patients compared with paired blood samples. Note that c3140 A->G nucleotide change causes H1047R mutation in protein. **B)** Bias in mutant allele detection in macrodactyly. The presence of the 3140A:G mutation in genomic DNA from dissociated nerve, adipose/fiberous tissue and blood from the same patient. **C)** expression of tissue markers, *MBP* for myleinated nerves, and *SMA* for vascular smooth muscle in excised macrodactyly tissue (scale bar, 20um). **D)** Pattern of expression of common regulators of overgrowth signaling pathways in macrodactyly. Sections of affected tissue show co-localization of growth related genes including PIK3CA to the myelin sheath of nerves (inset, scale bar, 5um) as well as smooth muscle and endothelial cells of the vasculature (inset; scale bar, 20um). Samples lightly counterstained with hematoxylin. *MBP*, myelin basic protein; *SMA*, smooth muscle myosin; *PTN*, pleiotrophin. *p-mTOR* is a phosphor-specific antibody.

### Growth related genes expressed in vasculature and nerves in macrodactyly tissue

PIK3CA is part of a growth program active in many cell types. We sought to understand PIK3CA regulation of overgrowth by identifying tissues in which it is regulated and that are particularly affected in the patients with macrodactyly. Using immunohistology of isolated tissue samples from macrodactyly and polydactyl control tissue we were able to localize PIK3CA protein to be highly regulated in the endothelial cells of arterioles as well as the myelinated cells of nerves (**Figure 2C**). Corresponding to these patterns of PIK3CA localization, we also found co-expression of active p-AKT and p-mTOR in these tissues as well, though not as prevalent as in nerve tissue (**Figure 2C**). In contrast, Pleiotrophin (PTN), a gene we found highly differentially expressed in macrodactyly (Lau et al., 2012), was predominantly expressed in smooth muscle of the vascular tissue. The observed patterns of expression in macrodactyly tissue were generally consistent with that seen in polydactyly samples which serve as a ‘nonaffected’, or wildtype PIK3CA, control tissue (**Supplementary Figure 1**).

### Identification of KNCH2 variant and correspondence with expression in affected tissues in patient population

Analysis of unique somatic or germline mutations within targeted growth related genes resulted in the identification of common shared mutations among the patient cohort. These mutations were confirmed via Sanger sequencing of the sample. One common mutation in the potassium channel, *KCNH2 (HERG/Kv11*.*1)* was quite prevalent. Given the frequency of the mutation and across tissue source, it is most likely germline. This variant is predicted to lead to a nonsynonymous substitution of threonine for lysine at position 897 of the channel. The K897T polymorphism has been observed previously and depending on context is thought to act as a dominant allele, modifying KCNH2 function in cardiac physiology as well as to modify the function of other potassium channel channels (Anderson Tawil Syndrome, KCNJ2 (Krych et al., 2017; Polyak et al., 2020); long QT syndrome, KCNQ1) (Ehrlich et al., 2004). In our cohort, we found 36% of patients (13/36) to be carriers of this mutation, with 2 patients being homozygous (**Table 2**). This frequency is consistent with sampling from patients with primarily European ancestry having estimates of minor allele frequencies of this variant (rs1805123) around 23-25% (dbSNP, GnomAD). The only patient carrying the K897T variant who did not identify as White/Caucasian, is from Hispanic background noted with relative high minor allele frequency of K897T (17%). Only two African American patients were identified in the cohort, both were wild type for KCNH2 consistent with low allele frequency in this population (4%, dbSNP, gnomAD).

Although analysis of *KCNH2* K897T allele frequency within human populations does not suggest pathogenicity, as homozygotes are retained within the general population (GnomAD) and well as observed in our patient cohort (**Table 2**), several lines of evidence suggest that the it could act to modify the effects of activated PIK3CA during growth of the distal appendages. Supporting a role for KCNH2 in orchestrating appendage development, modulation of the activity of potassium channel function is one of the primary means for coordinated overgrowth observed in fin size regulation of the zebrafish, causing normally patterned, albeit larger fins (Harris et al., 2021). Further, we recently identified upregulation of the zebrafish orthologue, *Kcnh2a*, as underlying increased size of the fins in the *longfin* mutant (Daane et al., 2021). Thus, as the K897T allele of *KCNH2* has been found to act in a modifying function in the heart and underlying developmental syndromes affecting face and limb (Crotti et al., 2005; Zhang et al., 2008), we reasoned that modulation of *KCNH2* may underly how coordinated overgrowth is balanced in macrodactyly, driven by oncogenic mutations in PIK3CA.

By immunohistology of affected patient digital overgrowth tissue and polydactyly controls, we assessed the expression of KCNH2 in relation to other known growth related genes. In polydactyly, KCNH2 is expressed in sensory nerves, predominantly detected in the perineurium and scattered sensory nerves (**Figure 3A**). Expression of KCNH2 in macrodactyly is broader in extent of the nervous tissues. For a large part this is coincident with enhanced perineurium in the affected nerves. In many cases expression was more broad, affecting epineurium and nerve associated cells as well. In the absence of KNCH2 897T mutations, we see varied expression (**Figure 3B**,**C**), whereas carriers of the *KCNH2* 897T mutation showed general misexpression (**Figure 3**). The expression patterns were reminiscent of those seen with PIK3CA and other growth regulators, suggesting activity in comparable tissues (**Figure 2**).

**Figure 3.**
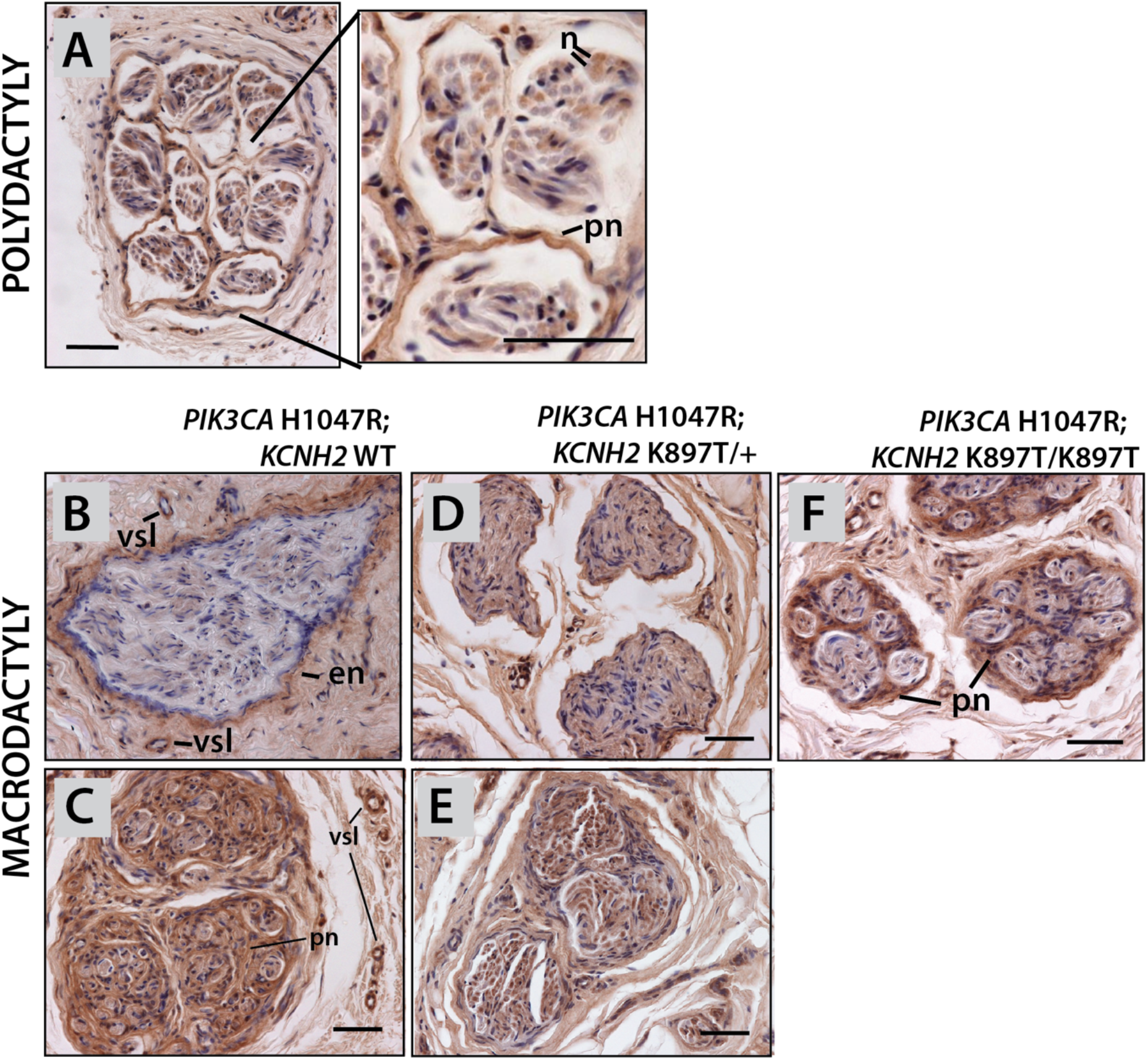
KCNH2 expression within affected tissue of macrodactyly. **A**. Anti-KCNH2 immunohistology shows KCNH is normally expressed in perineurium and can be seen in isolated cells within nerve bundle. B-F) Variable and expanded expression in macrodactyly tissue. **B-C)** KCNH2 expression observed in diverse pattern between patients positive for PIK3CA^H1047R^ without KCNH2^K897T^; Expression seen in epineurium in one case and also most tissues of the nerve in others. **D-E)** KCNH2 Expression in two patients harboring PIK3CA^H1047R^ and heterozygous KCNH2^K897T^. **F)** Prominent KCNH2 expression throughout perineurium of patient with PIK3CA^H1047R^; KCNH2^K897T/ K897T^ genotype. *n*, neuron; *en*, endoneurium; *pn*, perineurium; *vsl*, vessel. Scale bar equals 20um.

### Modeling macrodactyly in zebrafish

Prior work in mouse modeling PROS syndromes with activated PIK3CA in somatic clones has failed to recapitulate macrodactyly overgrowth phenotypes (Hare et al., 2015; Kinross et al., 2015). We tested sufficiency of human *PIK3CA* H1040T allele to drive overgrowth using the zebrafish model. The zebrafish provides an accessible experimental model for overgrowth syndromes as mutant clones can be easily generated in different and defined genetic backgrounds (**Figure 4A**). PIK3CA protein is highly conserved between human and zebrafish with over 92% identity of amino acid sequence, with high conservation within the region harboring the H1070R. We injected wildtype zebrafish eggs from strains (AB and Tu) with a transgene whereby *hPIK3CA*^H1070R^ is overexpressed via constitutive ubiquitin promoter (*Tg(ubi::hPIK3CA*^H1040R^). Injections of *ubi::PIK3CA*^H1040R^ surprisingly showed little effect overall on wildtype fish, showing no specific overgrowth of the fin (**Figure 4B**). Thus similar to previous mouse studies, activation of *PIK3CA* through the gain-of-function mutation H1040R is not sufficient to drive coordinated overgrowth of the extremities (limb or fin). However, when *ubi::hPIK3CA*^H1040R^ is injected into embryos having activated *kcnh2a* from the *lof* mutation, local overgrowths of the fin, or spikes, were observed at low frequency. These showed generally normal patterning and overgrowth is colocalized with clones expressing the transgene (**Figure 4C**). The growth effects extend beyond area of observed clones suggesting that the effect of *hPIK3CA*^H1040R^ expression may be non autonomous.

**Figure 4.**
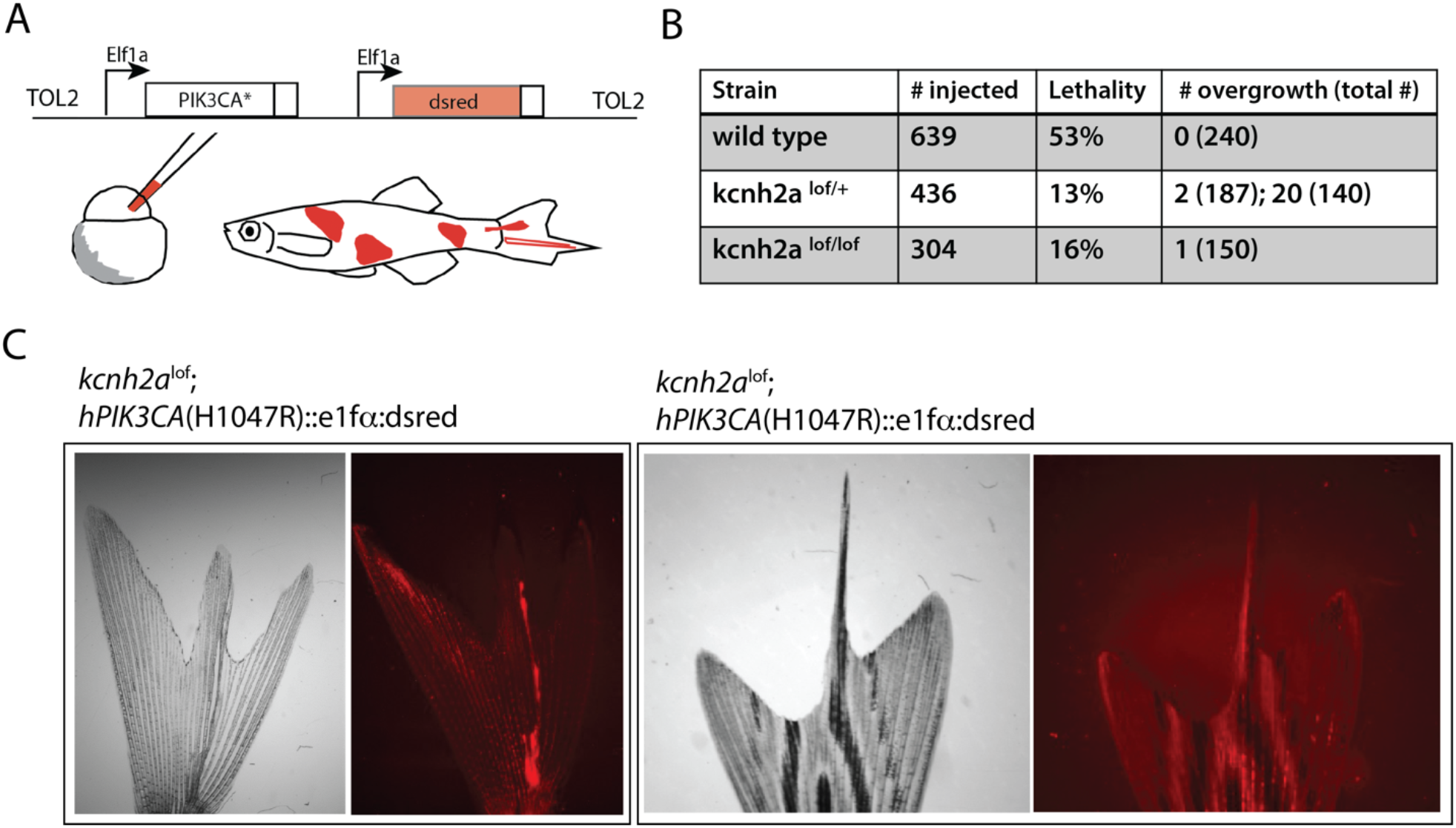
Genetic synergy between activated Kcnh2a and somatic clones driving h*PIKCA*^H1047R^ and phenocopy macrodactyly in the zebrafish. **A)** Experimental design to assess function of induced somatic mosaic clones of hPIK3CA^H1047R^. **B)** Summation of phenotypic consequences of hPIK3CA^H1047R^ induction on viability and formation of localized fin overgrowths. Dual data tabulated in case of separate experimentation. **C**). Morphology of caudal fin overgrowth observed and association with dsRed marked clone.

The effect of Kcnh2a in promoting patterned overgrowth in response to expression of activated *PIK3CA* could be direct acting in the fin itself or indirect, affecting growth more globally. To address indirect regulation of overgrowth potential we asked if there were early changes in mosaic PIK3CA embryos in the background of *lof*. Analysis of injected populations in wild-type or *kcnh2a* mutant background showed substantial reduced lethality in response to transformation by Pik3CA expression constructs (**Figure 4B**). The result of buffering by *kcnh2a* activity would presumably lead to persistence of higher clone density and number within juveniles and adults.

## DISCUSSION

One of the hallmarks of development is robustness – the suppression of variability such that with high probability similar, morphological and physiological systems are generated. This fact ensures functionality and homeostasis of the organism – when out of sync, dysmorphia, cancer and neoplastic growth can be the outcome. The coordination of tissue growth and patterning in cases of change highlights the involvement of such regulative processes in development. However, these regulatory processes and their roles in disease progression are not well understood.

We highlight the analysis of segmental overgrowth disorders in an effort to understand how such regulative action is controlled. We focus on the presentation of macrodactyly where unlike other segmental overgrowth disorders, growth is integrated, generating larger, albeit anatomically proportioned structure. Though dysmorphic growth can be observed, especially disproportionate dorsal/palmar growth, digits in macrodactyly patients are well patterned in comparison with other overgrowth disorders. We performed a systematic analysis of a novel cohort of macrodactyly patients. For a control, we compared genetic and structure of polydactylous tissue to parse out differences due to regulation in macrodactyly apart from ‘normal’ development of digital tissue. We find little clear anatomical (preaxial/central/postaxial) restriction or racial bias in our cohort: postaxial expression was less prevalent as previously noted in the literature (e.g.(Wu et al., 2020a)). White/Caucasian patients were more represented in our cohort (**Table 1**); however this is most likely this is a sampling artifact rather than bias in presentation.

We performed targeted sequencing of a panel of known oncogenes and included genes with evidence of growth regulation within these pathways. We specifically searched for mutations unique to affected tissue over those found germline (as identified in blood). We performed targeted sequencing on 18 patients. The vast majority of patients had somatic mutations within PIK3CA gene consistent with prior identification of associating *PIK3CA* mutations in macrodactyly and more broadly, PROS overgrowth disorders. Additionally, we detected common mutation in *PIK3CG*, S442Y, in 5/17 patients from our initial cohort, in individuals having *PIK3CA* mutations and without (**Table 2**). *PIK3CG* encodes a subunit of PIK3γ kinase. While PI3Kα and PI3Kβ are broadly expressed, PIK3γ is predominantly active in immune cells, vascular smooth muscle and cardiac cells (Perrotta et al., 2016). The S442Y mutation is predicted to be deleterious (Polyphen-2 0.984; SIFT) suggesting function of this change. Loss-of-function of *PIK3CG* in mice leads to defects in microglia (Hirsch et al., 2000). Thus, it is difficult to attribute growth function to this common identified variant within patients.

Of the identified mutations in *PIK3CA* across the larger cohort of patients, the H1047R allele was by far the most prevalent of known *PIK3CA* oncogenic mutations detected at low frequency (**table 2**). No other mutations within these gene sets were found and confirmed except for one patient having an *NFATC1* mutation. Thus, there is a particular role for *PIK3CA* mutations in driving macrodactyly even above other genetic factors in the same growth regulatory pathway, such as *AKT1, PTEN* and mTOR. Interestingly, this bias stands in stark contrast with the finding that *hPIK3CA*^H1047R^ is not sufficient to drive overgrowth that resembles macrodactyly. No mouse models of activated clonal expression of *PIK3CA*^H1047R^ have produced mosaic overgrowths of the limbs, nor have zebrafish overexpression analyses shown overgrowth of wildtype fins. However, it is clear from the mutational spectrum in patients that this mutation underlies this disorder. These results hint at genetic or environmental modifiers that promote coordinated overgrowth and manifestation of growth of activated PIK3CA somatic clone populations in the limb.

In our analysis, we identified prevalence of a common polymorphism mutation in KCNH2, K897T. Several lines of evidence suggest that this allele functions as a dominant modifier as it can buffer effects of disruption of the function of other key genes leading to arrythmia in LongQT syndrome(Crotti et al., 2005; Zhang et al., 2008). Our work in the zebrafish has shown that activation of Kcnh2a is sufficient to drive coordinated overgrowth leading to the hypothesis that this factor may potentiate inductive signals of activated PI3K signaling and lead to overgrowth. The K897T mutation itself does not have an obvious effect on phenotype or viability of humans as it shows high frequency in populations and no selection against the allele (gnomAD). Indeed, we find that Kcnh2 is expressed in comparable neural tissues as growth regulators including mTor, Akt and Pik3CA. It is noteworthy, however, that the effect on development leading to the overgrowth may extend beyond the immediate affected tissue we were able to study. Kcnh2 has been identified in the mouse apical ectodermal ridge which is required for proximal distal outgrowth of the limb in development (ref). Indeed, when clones expressing human PIK3CA^H1047R^ (hPIK3CA^H1047R^) are generated in zebrafish with increased Kcnh2a function, local fin overgrowths having normal coordinated growth are observed at low frequency. Although fin rays are not structurally similar to digits, their formation and outgrowth rely on similar developmental signals. This genetic combination thus provides the first evidence of recapitulation of macrodactyly-like growth in a vertebrate model, and suggests that Kcnh2 activity, or the developmental consequences of enhanced Kcnh2 activity, can potentiate overgrowth due to driving *PIK3CA* mutations.

The potassium channel KCNH2 is known to be modulated by a host of intracellular regulators such as Protein kinase B and Akt. Interestingly, PI3K influences the activity of KCNH2 and other channels by affecting the levels of PIP2, which can indirectly or directly modulate ion-channel function (27). Finally, membrane polarization caused by activated channels can indirectly affect PIP2 levels and downstream signaling by altering intracellular Ca2+ levels. As such, Kcnh2 is poised to act within a common integrated pathway as PI3K signaling in regulation of growth, linking bioelectric cues to canonical signaling pathways. There is also evidence that Kcnh2 activity is stimulated by thyroid hormone through PP5/ RacGTPase signaling downstream of Pi3K (Gentile et al., 2008; Storey et al., 2006; Storey et al., 2002). The regulation of KCNH2 activity with thyroid hormone provides an interesting link to the timing and local regulation of overgrowth in macrodactyly that deserves further investigation.

In an insightful early paper detailing sensory nerve involvement in the etiology of macrodactyly, B. H. Moore postulated on the causes of macrodactyly as an interplay between permissive factors and ones limiting or directing action of these growth impulses into a ‘normal’ pattern (Moore, 1942). As he noted, loss of this regulative mechanism would lead to abnormal growth; however in the same logic, activation or maintenance of activity might bolster extended patterned growth given an underlying inductive signal. Here we provide the first model of macrodactyly through genetic interaction between Pik3ca and Kcnh2 in the zebrafish fin. A genetic association between the KCNH2^897T^ and PIK3CA^H1047R^ mutations is not apparent in our patient cohort. However, as we have found that increased activity of Kcnh2a underlies overgrowth potential, non-mutant individuals may involve modulation of KCNH2 activity by other means than activating mutation. Broader analysis of co-expression profiles within cells of affected tissue in macrodactyly may provide important insight into the signature of activation and commonalities with conditions of active KCNH2 or PIK3CA function. As KCNH2/HERG is a common target of small molecule regulators, the modulation of this signaling activity may provide a novel means of shaping overgrowth especially in cases of progressive growth which can be debilitating.

## METHODS

### Clinical Sample collection

After being granted approval from our institutional review board, we prospectively received informed, written consent to collect discarded tissue from patients undergoing surgery for macrodactyly or polydactyly from December 2012 through April 2017 at Boston Children’s Hospital. We also obtained additional macrodactyly and polydactyly samples with a waiver of informed consent by retrospectively querying the pathology database located at Boston Children’s Hospital for patients who had undergone previous surgical correction of macrodactyly or polydactyly.

### Clinical data management and statistical analysis

All statistical analyses were performed using IBM SPSS Version 24 (IBM Corp., Armonk, N.Y.). Frequencies were tabulated for demographics, clinical data, and mutation types. Lipomatous macrodactyly was defined as fatty overgrowth disproportionate to the size of the digit, and nerve-territory macrodactyly was defined as overgrowth of the nerve disproportionate to other tissues in the digit. Macrodactyly was classified as secondary if it presented as a direct result of another underlying condition. Medians were calculated for continuous variables, as necessary. Fischer exact tests were used to compare overall mutation status, and mutation type by location, age, sex, and race. All diagnoses were made using physical exam and clinical presentation. Unless otherwise stated, race, age, and mutation type were respectively dichotomized as follows: white, non-Hispanic versus all other races/ethnicities, and under 1.5 years at initial surgery versus over 1.5 years at initial surgery. A threshold of 10% missing data was used, and results were considered statistically significant when *p* < 0.05.

### DNA sequencing and exome capture

Somatic mutations in overgrowth disorders may only be present in a small subset and can be difficult to detect without high sequencing read coverage. We performed a targeted sequencing approach to recover high sequencing-read depth across a panel of 24 overgrowth-related genes, including genes associated with human overgrowth disorders (*AKT1, AKT2, AKT3, PIK3CA, PIK3CG, PIK3CB, PIK3R1, PIK3R2*), zebrafish appendage overgrowth mutants (*KCNK5, KCNH2, KCNH1, SLC12A7*), tumor suppressors associated with nerves (*NF1, NF2*) genes that function in calcium signaling (FKBP1A, Calcineurin (*PPP3CA, PPP3CB, PPP3CC*), and *NFATC1*), and genes that are upregulated in patients with macrodactyly (*IGF1, PTN, MDK, NOS3*)^1^. We designed a custom Agilent SureSelect sequence capture microarray composed of 100,000, 60mer oligo probes that are spaced every 1 bp across the targeted genes and each probe repeated 6-7 times.

Genomic DNA was extracted with the Qiagen DNeasy Blood & Tissue extraction kit. A total of 3-5μg DNA from each tissue sample was sheared to an average size of 200bp using a Covaris E220 ultrasonicator (duty cycle, 10%; intensity, 5; cycles/burst, 200; time, 180 seconds; temperature, 8°C). DNA libraries were blunt-ended, 5’ phosphorylated, A-tailed, adaptor ligated and hybridized to the capture array at 65°C as previously described in Bowen *et al* (Bowen et al., 2011).Both pre- and post-capture libraries were amplified with Phusion High-Fidelity DNA Polymerase (NEB # M0530). The 19 samples were split between two capture arrays and sequenced at 100bp single end reads on one lane of Illumina HiSeq 2000.

Sequencing reads were barcode sorted, adapter trimmed and aligned to the human genome (hg19) using Novoalign (http://novocraft.com). PCR duplicates were removing using the Genome Analysis Toolkit (GATK) (parameters ‘MarkDuplicates.jar ASSUME_SORTED=true REMOVE_DUPLICATES=true’).

We used a threshold of 4% allele frequency and at least two sequencing reads before considering putative somatic mutations. We further excluded candidate somatic mutations that were present within the larger population using the NHLBI Exome Variant Server (EVS). The effect of coding variants was assessed using SNPEff (GRCh37.68)^3^.

### PCR and Colony Sanger Sequencing of Patient Samples

Exome sequencing results were confirmed and expanded upon by using Sanger sequencing. DNA was purified from patient tissue using TRI Reagent (Sigma #1001877531) or DNeasy Blood and Tissue Kit (QIAgen #69506) according to the manufacturer’s instructions.

*PIK3CA* exons 21 or 8 were sequenced for the mutations H1047R or C420R respectively. Primer sequences for PCR of the putative exons containing mutations are as follows. Exon 21 F: TTGATGACATTGCATACATTCG. Exon 21 R: GGAATCCAGAGTGAGCTTTCA. Exon 8 F: GTAAAACGACGGCCAGTTCCCATTATTATAGAGATGATTGTTGA. Exon 8 R: AACAGCTATGACCATGTGGATTTGATCCAGTAACACCA.

*KCNH2* exon 11 was PCR amplified for the K897T mutation using the following primer set. Exon 11 F: AGGTCTGAGGCCTGGGTAAA. Exon 11 R: CGGAGTTAGAGGGTGGCTTC. After amplification, PCR products were ligated into either pGEM T-Easy (Promega #A137A) or pJET1.2 (Thermo Scientific #K1231), and transformed into TOP10 bacteria. DNA was mini-prepped from between 15-30 colonies for each patient sample and sent to Beckman Coulter for Sanger sequencing.

### Histology and immunohistochemistry

Tissue samples were fixed overnight at 4°C in 4% paraformaldehyde. Fixative was removed through three washes of phosphate buffered saline (PBS) prior to a dehydration series of 30/50/75/100% ethanol. Samples were then rinsed in CitriSolv (Decon Labs #1601H), embedded in paraffin wax and stored at 4°C. Tissues sections were deparaffinized in an ethanol series and rehydrated in distilled water (dH_2_O). Endogenous alkaline phosphatase activity was blocked in 1% hydrogen peroxide (H_2_O_2_) for 15 minutes and rinsed for 5 minutes in PBS. We performed antigen retrieval by boiling slides in 10 mM sodium citrate (pH 6.0) for 30 minutes at 90°C and cooled to room temperature. Slides were antigen blocked for 2 hours in PBST (PBS + 5% heat inactivated goat serum + 0.1% Tween). Primary antibodies were incubated for 1 hour at room temperature. Primary antibody and concentrations: p-AKT (1:50, Cell Signaling #4060), p-MTOR (1:100, Cell Signaling #2976), p-PIK3CA (1:50, Cell Signaling #4249), and KCNH2 (1:400, ThermoFisher Scientific #PA3-860). Primary antibody was rinsed from the slides with PBST (5 minutes) and then incubated at room temperature for 40 minutes with secondary antibody diluted in PBST (1:500 Biotin-conjugated Goat anti-Rabbit IgG; ThermoFisher #31820). Following secondary antibody incubation, slides were exposed to ABC reagent (Vectastain #PK-7100) for 20 minutes, followed by 30 seconds in 1% Tween solution and then 30 seconds in dH_2_O. The slides were then exposed in diaminobenzidine (DAB, Sigma #D0426) for up to 15 minutes. After exposure, slides were rinsed in dH_2_O, dipped in hematoxylin for up to 20 seconds, rinsed in dH_2_O again and dehydrated into an ethanol series to xylene for cover-slipping and imaging. Polydactyly digital nerve samples: PS007, PS013. Macrodactyly digital nerve samples: PS006, PS011.

### Overexpression of Mutant Human PIK3CA in Danio rerio

The human *PIK3CA* gene was cloned into a Tol2 Invitrogen Gateway destination vector 394 with an EF1a promoter and an dsRed marker. DNA was injected into one-celled AB or LOF embryos at a concentration of 5-15 ng/ul. Lethality was assessed over a 1-3 day period, and dsRed positive embryos were allowed to grow to adulthood (3-4 months old) and then examined for fin overgrowths.

## Data Availability

All data produced in the present study are available upon reasonable request to the authors

## Acknowledgements

The authors would like to acknowledge all patient and patient families who agreed to participate in this study. This work was partially supported by NIH R01HD084985 and BSF US-Israel BSF (grant no. 2017204) to MPH.

---

**Supplementary Figure 1.**
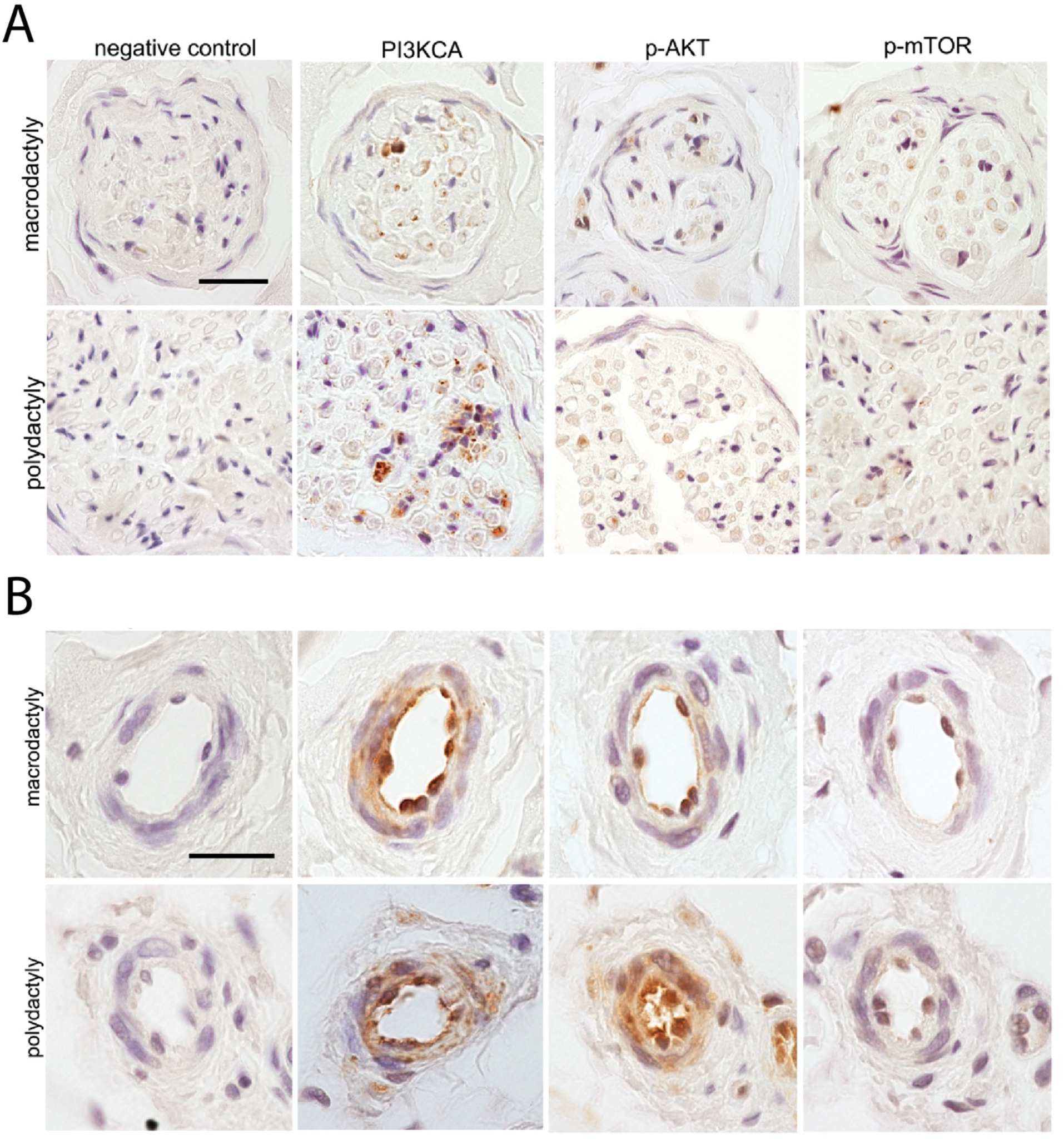
Immunohistochemistry of growth related genes in **A**) nerves and **B**) vascular tissues in macrodactyly and polydactyly patients. Scale bar equals 20um.

**Supplementary Table 1.**

| Patient | Sex | Race | phenotype oriented | Isolated/ Secondary | Hand / Foot | Side | Affected Digits | Territory |
| --- | --- | --- | --- | --- | --- | --- | --- | --- |
| 1 | F | White | Nerve-territory | Isolated | Hand | L | 1,2 | Preaxial |
| 2 | F | White | Lipomatous* | Isolated | Foot | L | 2,3 | Preaxial |
| 3 | M | White | Lipomatous | Secondary | Foot | L | 1,2,3 | Preaxial |
| 4 | M | White | Nerve-territory | Isolated | Hand | L | 1,2,3 | Preaxial |
| 5 | M | White | Nerve-territory | Isolated | Hand | L | 1,2 | Preaxial |
| 6 | M | Hispanic | Lipomatous | Isolated | Foot | L | 1,2,3 | Preaxial |
| 7 | F | Other | Lipomatous | Isolated | Foot | L | 1,2 | Preaxial |
| 8 | M | Unknown | Lipomatous | Isolated | Foot | L | 1,2,3 | Preaxial |
| 9 | M | Afri-Am. | Nerve-territory | Isolated | Hand | L | 1,2 | Preaxial |
| 10 | F | White | Nerve-territory | Isolated | Hand | L | 1,2 | Preaxial |
| 11 | F | Unknown | Nerve-territory | Isolated | Foot | R | 2 | Preaxial |
| 12 | F | White | Lipomatous | Isolated | Foot | L | 1,2 | Preaxial |
| 13 | M | White | Nerve-territory | Isolated | Foot | R | 1,2 | Preaxial |
| 14 | F | White | Lipomatous | Isolated | Foot | R | 1,2 | Preaxial |
| 15 | F | White | Lipomatous | Isolated | Foot | L | 1,2 | Preaxial |
| 16 | F | White | Nerve-territory | Isolated | Foot | L | 1,2 | Preaxial |
| 17 | F | White | Lipomatous* | Secondary | Hand | R | 1,2,3 | Preaxial |
| 18 | M | Other | Nerve-territory | Isolated | Foot | R | 2,3 | Central |
| 19 | F | Other | Lipomatous | Isolated | Hand | L | 2,3 | Central |
| 20 | F | Unknown | Lipomatous | Isolated | Hand | R | 3 | Central |
| 21 | M | Asian | Lipomatous | Isolated | Hand | L | 3,4 | Central |
| 22 | M | Hispanic | Lipomatous | Isolated | Foot | R | 3,4 | Central |
| 23 | M | White | Lipomatous | Isolated | Hand | R | 2,3,4 | Central |
| 24 | M | Unknown | Nerve-territory | Isolated | Hand | L | 3,4 | Central |
| 25 | M | Afri-Am. | Nerve-territory | Isolated | Foot | R | 2,3 | Central |
| 26 | F | White | Lipomatous | Secondary | Foot | R | 3,4 | Central |
| 27 | M | White | Nerve-territory | Isolated | N/A | L | 4 | Central |
| 28 | M | Other | Nerve-territory | Isolated | Hand | L | 3,4 | Central |
| 29 | F | White | Lipomatous | Isolated | Foot | L | 2,3,4 | Central |
| 30 | M | White | Lipomatous | Isolated | Hand | L | 2,3 | Central |
| 31 | M | White | Nerve-territory | Isolated | Hand | L | 3,4 | Central |
| 32 | F | Hispanic | Nerve-territory | Isolated | Hand | R | 2,3 | Central |
| 33 | F | White | Nerve-territory | Isolated | Hand | L | 3,4,5 | Postaxial |
| 34 | F | White | Nerve-territory | Isolated | Hand | L | 4,5 | Postaxial |
| 35 | F | White | Lipomatous* | Secondary | Foot | L | 3,4,5 | Postaxial |
| 36 | M | Other | Lipomatous | Isolated | Foot | L | 3,4,5 | Postaxial |

\* Hemi-hypertrophy as secondary diagnosis

**Supplementary Table 2.** Targeted capture array gene list.

| Gene | Avg. Depth | Avg coverage >2 reads | Avg coverage >10 reads | Confirmed mutation* | Freq of mutations in macrodactyly patients |
| --- | --- | --- | --- | --- | --- |
| *AKT1* | 43.1 | 100.0% | 99.9% |  |  |
| *AKT2* | 44.5 | 100.0% | 99.3% |  |  |
| *AKT3* | 22.4 | 98.0% | 85.9% |  |  |
| *FKBP1A* | 7.4 | 59.5% | 33.2% |  |  |
| *IGF1* | 35.5 | 100.0% | 96.1% |  |  |
| *KCNH1* | 34.7 | 99.9% | 94.4% |  |  |
| *KCNH2* | 27.5 | 91.8% | 74.6% | K899T |  |
| *KCNK5* | 36.0 | 100.0% | 99.4% |  |  |
| *NF1* | 21.9 | 95.4% | 78.5% |  |  |
| *NF2* | 24.2 | 98.4% | 81.3% |  |  |
| *NFATC1* | 23.5 | 97.9% | 82.6% | S299L | 1/18 |
| *MDK* | 29.0 | 100.0% | 97.9% |  |  |
| *MTOR* | 32.2 | 99.9% | 96.0% |  |  |
| *NOS3 (eNOS)* | 25.9 | 95.6% | 84.9% |  |  |
| *PIK3CA* | 27.8 | 93.2% | 87.8% | H1047R; E542K | 14/18 |
| *PIK3CB* | 24.1 | 99.8% | 88.8% |  |  |
| *PIK3CG* | 36.7 | 99.9% | 97.9% | S442Y | 5/17 |
| *PIK3R1* | 24.7 | 99.7% | 85.0% |  |  |
| *PIK3R2* | 26.9 | 87.4% | 74.7% |  |  |
| *PIK3R3* | 29.0 | 99.7% | 91.1% |  |  |
| *PPP3CA* | 23.4 | 98.5% | 83.6% |  |  |
| *PPP3CB* | 17.8 | 95.9% | 72.6% |  |  |
| *PPP3CC* | 24.6 | 100.0% | 88.1% |  |  |
| *PTEN* | 10.9 | 79.2% | 47.1% |  |  |
| *PTN* | 25.4 | 99.5% | 84.4% |  |  |
| *SLC12A7* | 30.1 | 97.6% | 94.3% |  |  |
| **average** | **27.2** | **95.6%** | **84.6%** |  |  |

